# Stereotactic Ablative Radiotherapy for the Comprehensive Treatment of Oligometastatic Cancers: Long-Term Results of the SABR-COMET Phase II Randomized Trial

**DOI:** 10.1101/2020.03.26.20044305

**Authors:** David A. Palma, Robert Olson, Stephen Harrow, Stewart Gaede, Alexander V. Louie, Cornelis Haasbeek, Liam Mulroy, Michael Lock, George B. Rodrigues, Brian P. Yaremko, Devin Schellenberg, Belal Ahmad, Sashendra Senthi, Anand Swaminath, Neil Kopek, Mitchell Liu, Karen Moore, Suzanne Currie, Roel Schlijper, Glenn S. Bauman, Joanna Laba, X. Melody Qu, Andrew Warner, Suresh Senan

**Author notes:** **Corresponding Author:** Dr. David Palma, MD, PhD, London Health Sciences Centre, 800 Commissioners Rd. E., P.O. Box 5010, STN B, London, Ontario, Canada N6A 5W9.

## Abstract

**Purpose:** The oligometastatic paradigm hypothesizes that patients with a limited number of metastases may achieve long-term disease control, or even cure, if all sites of disease can be ablated. However, long-term randomized data testing this paradigm are lacking.

**Methods:** We enrolled patients with a controlled primary malignancy and 1-5 metastatic lesions, with all metastases amenable to stereotactic ablative radiotherapy (SABR). We stratified by the number of metastases (1-3 vs. 4-5) and randomized in a 1:2 ratio between palliative standard of care (SOC) treatments (Arm 1) vs. SOC plus SABR (Arm 2). We employed a randomized phase II screening design with a primary endpoint of overall survival (OS), using an alpha of 0.20 (wherein a p-value <0.20 indicates a positive trial). Secondary endpoints included progression-free survival (PFS), toxicity, and quality of life (QOL). Herein we present long-term outcomes from the trial.

**Results:** Between 2012 and 2016, 99 patients were randomized (33 in Arm 1, 66 in Arm 2) at 10 centres internationally. Median age was 68 (range 43-89) and the majority (n=59; 60%) were male. The most common primary tumor types were breast (n=18), lung (n=18), colorectal (n=18), and prostate (n=16). Median follow-up was 51 months. Five-year OS was 17.7% in Arm 1 (95% confidence interval [CI]: 6-34%) vs. 42.3% in Arm 2 (95% CI: 28-56%; stratified log-rank p=0.006). Five-year PFS was ‘not reached’ in Arm 1 (3.2% [95% CI: 0-14%] at 4-years with last patient censored) and was 17.3% (95% CI: 8-30%) in Arm 2 (p=0.001). There were no new grade 2-5 adverse events and no differences in QOL between arms.

**Conclusions:** With extended follow-up, the impact of SABR on OS was larger in magnitude than in the initial analysis, and durable over time. There were no new safety signals, and SABR had no detrimental impact on QOL. (NCT01446744)

**Funding:** Ontario Institute for Cancer Research and London Regional Cancer Program Catalyst Grant

## Introduction

It has been hypothesized for nearly a century that patients with a small burden of metastatic disease can benefit from ablation of all metastases,^1^ with some achieving long-term disease control or even cure. Although surgery was historically the primary modality used to ablate metastases,^2^ newer and less-invasive modalities are now available, including stereotactic ablative radiotherapy (SABR).^3^ Although the clinical use of surgery and SABR has increased rapidly in recent decades,^3,4^ randomized data proving the existence of the oligometastatic state have been lacking.^5^

Pre-clinical and translational studies provide evidence in support of the oligometastatic hypothesis.^6^ The development of metastases requires a series of key steps, known as the invasion-metastasis cascade.^7^ The cascade includes steps leading to tumor cell intravasation into the circulatory system, with subsequent hematogeneous dissemination and extravasation, followed by survival and colonization in a distant organ. The large majority of cells that reach a distant organ either die or enter dormancy, whereas only a small percentage proliferate to develop into metastases.^6,7^ Phylogenetic analyses of primary tumors and metastases using next-generation sequencing have allowed for the creation of timelines of metastasis development.^8-10^ In some patients, a solitary metastasis can be present for years as a single site of disease, but then subsequently seed further widespread metastases thereafter.^8-10^ Metastases can also re-seed the primary tumor.^11^

The biological evidence is supported by numerous single-arm studies testing ablative therapies in patients with oligometastases.^5^ Such studies have often demonstrated ‘better-than-expected’ long-term survivals for a population of patients with metastatic disease. However, conclusions from single-arm studies are limited by a lack of a control arm, leading to uncertainties as to whether long-term survivals reported are due to the ablative therapies themselves, or merely due to the selection of fit patients with slow-growing indolent disease.^5^ This debate has resulted in substantial international variation in the patterns of practice in treating patients with oligometastases.^12^

There is now supportive data from randomized phase II studies testing the impact of ablative therapies on overall survival (OS)^13,14^ or on surrogate endpoints such as progression-free survival (PFS).^13-18^ One of these, titled *Stereotactic Ablative Radiotherapy for the Comprehensive Treatment of Oligometastases* (SABR-COMET), assessed the impact of SABR on OS in patients with a controlled primary tumor and 1-5 metastatic lesions. The initial report of SABR-COMET demonstrated a 13-month improvement in median OS, the primary endpoint, after a median follow-up of 28 months.^14^ However, due to the larger-than-expected number of patients achieving 5-year survival, the SABR-COMET protocol was modified to extend follow-up beyond 5-years, in order to capture long-term outcomes. Herein we report the extended outcomes of the trial, more than 40 months after completion of accrual.

## Methods

### Study Design

SABR-COMET was an open-label phase II randomized international study that enrolled patients from 10 centers. Appropriate regulatory approval, including ethics approval, was obtained in all jurisdictions. The trial was registered (NCT01446744) prior to activation. Since the trial details and statistical analyses have been published in detail,^14,19^ a short synopsis is provided here.

### Participants

The main inclusion requirements were age ≥ 18 years, Eastern Cooperative Oncology Group performance score 0-1, a life expectancy ≥ 6 months or more, and patients were required to have 1-5 metastases and a controlled primary tumor. All metastatic lesions had to be eligible for SABR in accordance with protocol specified dose-constraints. The main exclusion criteria included serious medical comorbidities prohibiting radiotherapy, prior radiotherapy to a site requiring treatment, malignant pleural effusion, tumors in close proximity to the spinal cord (within 3 mm), and brain metastasis requiring surgical decompression. All patients provided written informed consent.

### Randomization and Masking

Patients were randomized using a computer-generated randomization list with permuted blocks of nine, after stratification by the number of metastases (1-3 vs. 4-5). There was no blinding of patients or physicians.

### Procedures

In the control arm, standard palliative radiotherapy was delivered with the goal of alleviating symptoms or preventing complications, with recommended doses ranging from 8 Gy in 1 fraction to 30 Gy in 10 fractions. In the SABR arm, patients received SABR to all sites of metastatic disease. A full table of allowable SABR doses is provided in the protocol (Supplementary Appendix). Patients in the SABR arm who subsequently developed new metastases were eligible for further SABR if feasible. In both arms, palliative standard of care (SOC) systemic therapy was recommended as indicated, using a pragmatic approach wherein the choice of systemic agents was at the discretion of the medical oncologist. Any further palliative systemic therapy or palliative radiation therapy after progression were at the discretion of the treating physicians.

Patients were seen in follow-up every three months after randomization in years 1-2, and every six months until year 5, with regular imaging as outlined in the protocol. The trial was amended in October 2016 to continue annual visits until year 10.

### Outcomes

The primary endpoint was OS, and secondary endpoints included quality of life (QOL), assessed with the Functional Assessment of Cancer Therapy: General (FACT-G); toxicity, based on the National Cancer Institute Common Toxicity Criteria (NCI-CTC) version 4; progression-free survival (PFS); lesional control rate; and the number of cycles of further chemotherapy/systemic therapy. This latter endpoint was not easily ascertainable because of patients receiving palliative systemic therapy at other centres, and was therefore reported as a binary variable (i.e. further systemic therapy received: yes vs. no) in the primary analysis and again here. An additional *post-hoc* endpoint was analyzed here: time to development of new metastases, defined as time from randomization to development of new metastatic lesions, treating death from any cause as a competing event. To address the possible imbalance arising from the distribution of prostate patients between the two arms, a *post-hoc* sensitivity analysis was performed to examine the impact of SABR on OS after excluding all prostate patients.

### Statistical Methods

SABR-COMET employed a randomized phase II screening design^14^, using a two-sided alpha of 0.20 and a power of 80%. In this approach, the alpha is set higher than the 0.05 level used in phase III trials, recognizing that even if the phase II trial is positive (i.e. if the p-value for the primary endpoint is below 0.20), such a positive result is not usually considered definitive proof without a subsequent phase III trial. However, a finding with p<0.005 in a phase II screening trial may be considered definitive.^20^

All analyses were based on the intention-to-treat principle. OS and PFS were calculated using the Kaplan-Meier estimates and differences were compared using the stratified log-rank test (adjusting for stratification). Hazard ratios (HRs) were calculated using Cox proportional hazards regression adjusted for stratification. Quality of life was measured using FACT-G scores, with differences between groups over time compared using linear mixed modelling (with time and treatment arm as fixed effects and patient number as random effect). Differences in rates of grade 2 or higher toxicity and in receipt of systemic therapy were compared using the Chi-square test or Fisher’s Exact test as appropriate. Time to development of new metastases was estimated using cumulative incidence functions with death considered a competing event, and differences compared using the stratified Gray’s test (adjusting for stratification). Statistical analysis was performed using SAS version 9.4 software (SAS Institute, Cary NC, USA), using two-sided statistical testing at the 0.05 significance level.

The trial closed in August 2016, and after a year of follow-up and time to resolve data queries, the dataset was locked for the previously published primary analyses on January 18, 2018. Data collection continued thereafter, and the dataset was locked for this long-term analysis on January 30, 2020. The first author (DP) and statistician (AW) had full access to the data, vouch for the integrity of the data and the adherence to the study protocol, and are responsible for the decision to submit the manuscript.

## Results

Between February 2012 and August 2016, 99 patients were enrolled at 10 centres; 33 in the control arm and 66 in the SABR arm (Figure 1). Baseline characteristics are shown in Table 1. Notably, the SABR group contained the preponderance of patients with prostate cancer and all the patients with five metastases.

**Table 1.** Baseline characteristics, reproduced from original publication.^14^

| **Characteristic** | **Control Arm**<br>**(n=33)** | **SABR Arm**<br>**(n=66)** |
| --- | --- | --- |
| **Age – median (IQR)** | 69 (64, 75) | 67 (59, 74) |
| **Sex – n(%)** |  |  |
| Male | 19 (58) | 40 (61) |
| Female | 14 (42) | 26 (39) |
| **Site of Original Primary Tumor – n(%)** |  |  |
| Breast | 5 (15) | 13 (20) |
| Colorectal | 9 (27) | 9 (14) |
| Lung | 6 (18) | 12 (18) |
| Prostate | 2 (6) | 14 (21) |
| Other | 11 (33) | 18 (27) |
| **Time from Diagnosis of Primary**<br>**Tumor to Randomization (years) –**<br>**median (IQR)** | 2.3 (1.3, 4.5) | 2.4 (1.6, 5.3) |
| **Number of Metastases – n(%)** |  |  |
| 1 | 12 (36) | 30 (46) |
| 2 | 13 (40) | 19 (29) |
| 3 | 6 (18) | 12 (18) |
| 4 | 2 (6) | 2 (3) |
| 5 | 0 (0) | 3 (5) |
| **Location of Metastases (n=191 lesions) – n(%)** |  |  |
| Adrenal | 2 (3) | 7 (6) |
| Bone | 20 (31) | 45 (35) |
| Liver | 3 (5) | 16 (13) |
| Lung | 34 (53) | 55 (43) |
| Other* | 5 (8) | 4 (3) |

\*Other includes brain (n=4 lesions), lymph nodes (n=4 lesions), and para-renal (n=1 lesion).
SABR – Stereotactic ablative radiotherapy; IQR – interquartile range.

**Figure 1.**
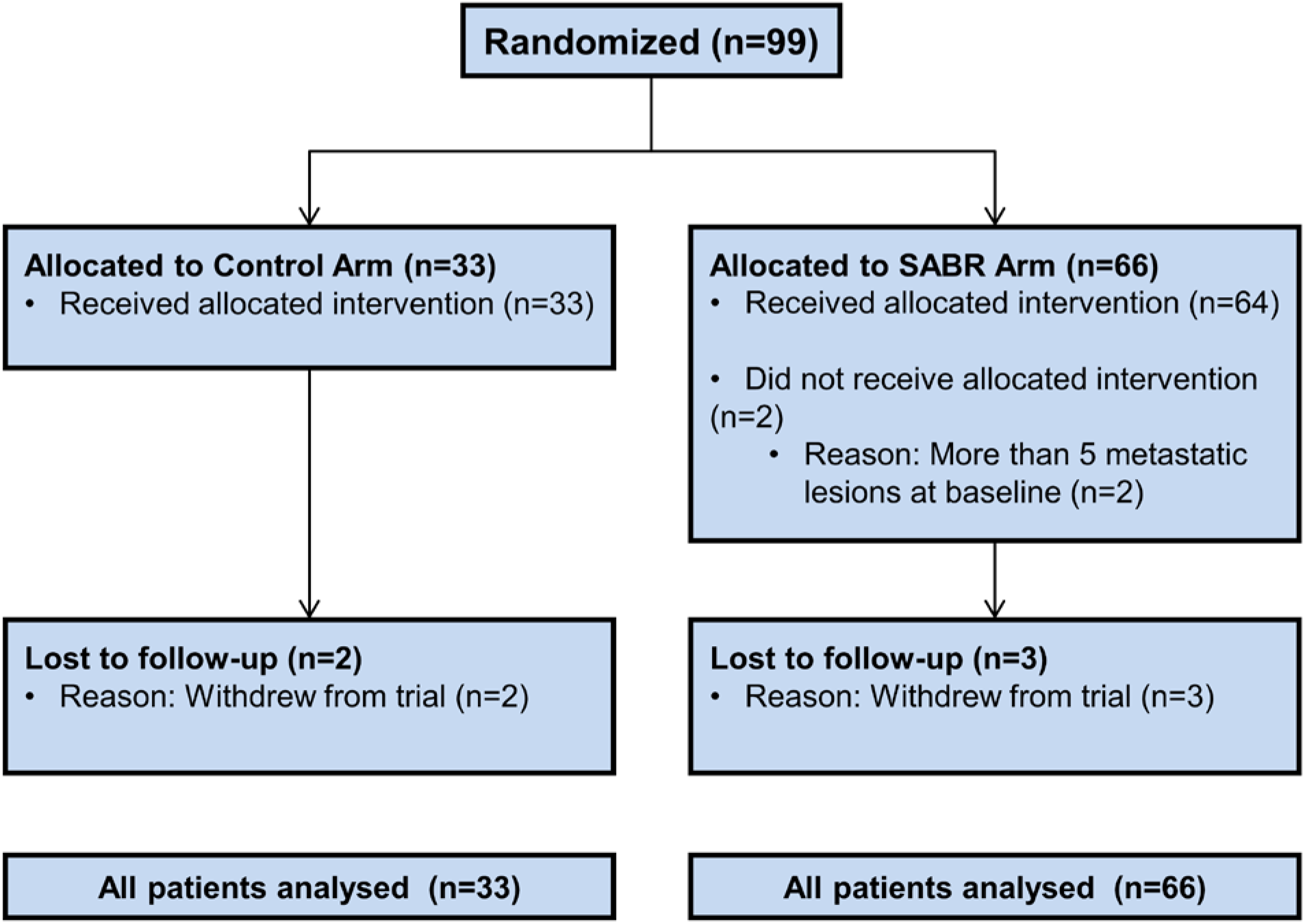
CONSORT Diagram.

After randomization, 57/99 patients (58%) received palliative systemic therapy, and 39/99 (39%) received palliative radiotherapy. Utilization of palliative radiotherapy was higher in the control arm (delivered to 23/33 patients [70%]) compared to the SABR arm (delivered to 16/66 patients [24%], p<0.001). There were no differences between arms in use of systemic therapy (21/33 [64%] vs. 36/66 [55%] respectively, p=0.39). Since the original report, one patient in the control arm received curative-intent SABR for a solitary liver lesion that had initially responded to targeted therapy but progressed with no new sites of disease. This patient remains alive and free of disease and is analyzed on the control arm. Nine patients in the SABR arm received salvage SABR for new metastases, including 30% of patients (3/10) surviving beyond 5-years.

The median follow-up was 51 months (95% confidence interval [CI]: 46-58 months). The primary outcome event, death from any cause, occurred in 24/33 patients (73%) in the control arm and 35/66 patients (53%) in the SABR arm. Median OS was 28 months in the control arm (95% CI: 18-39 months) vs. 50 months in the SABR arm (95% CI: 29-83 months; stratified log-rank test: p=0.006; hazard ratio [HR]: 0.47; 95% CI: 0.27-0.81; Figure 2a). Five-year OS rates were 17.7% (95% CI: 6-34%) vs. 42.3% (95% CI: 28-56%), respectively. A *post-hoc* sensitivity analysis excluding prostate patients was consistent with a treatment benefit with 5-year OS rates of 16.2% (95% CI: 5-32%) vs. 33.1% (95% CI: 20-47%) respectively (stratified log-rank test: p=0.085).

**Figure 2.**
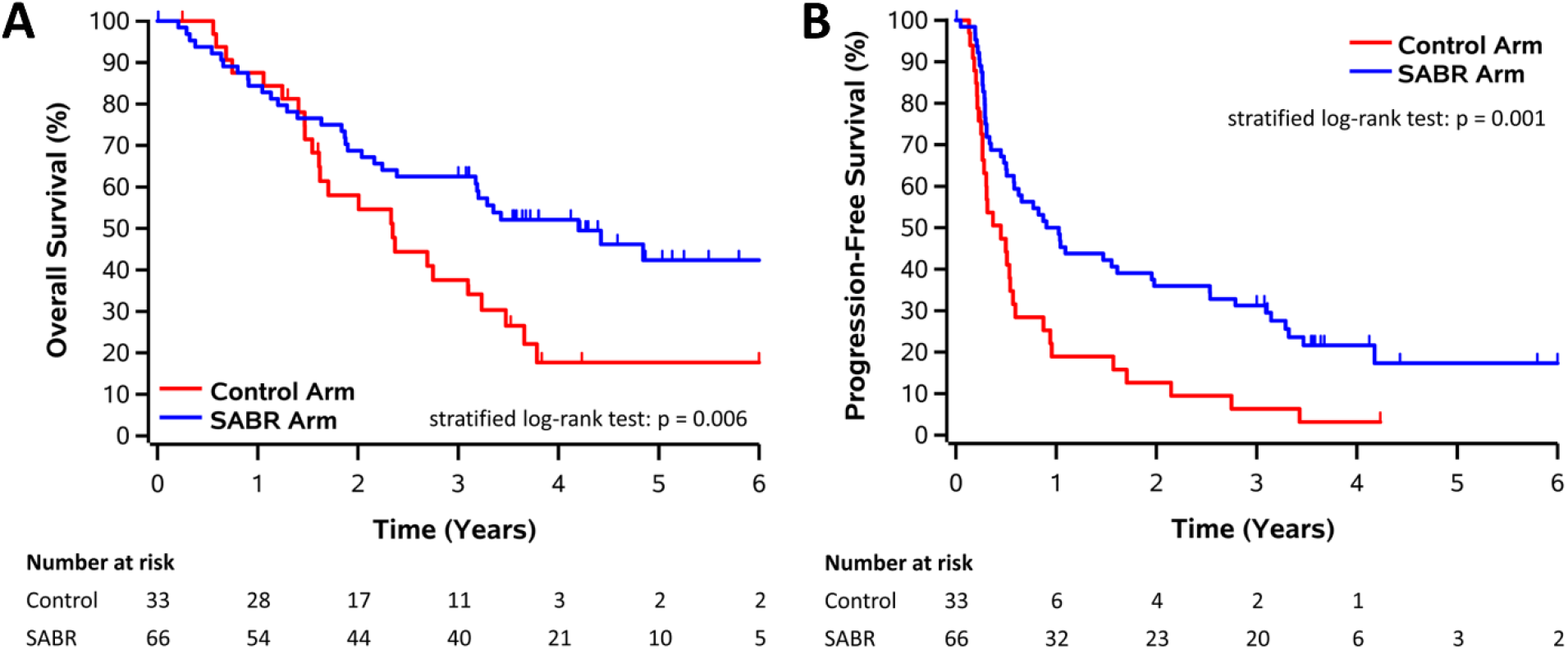
Kaplan-Meier plots for (A) overall survival (B) progression-free survival.

Progression events occurred in 74 patients, 29/33 patients (88%) in the control arm and 45/66 patients (68%) in the SABR arm. No patient in the control arm survived 5 years without progression (PFS was 3.2% [95% CI: 0-14%] at 4-years with last patient censored), whereas 5-year PFS was 17.3% (95% CI: 8-30%) in the SABR arm (stratified log-rank test: p=0.001; HR: 0.48, 95% CI: 0.31-0.76; Figure 2b).

The overall long-term lesional control (LC) rate, defined as the absence of progression in the lesions initially present at randomization based on RECIST 1.1 criteria, was 46% (26/57 assessable lesions) in the control arm and 63% (65/104 assessable lesions) in the SABR arm (p=0.039), corresponding to an absolute increase of 17% (95% CI: 1-33%). After SABR, there were significant differences in LC rates based on lesion location (LC rates: adrenal 100%, bone 72%, lung 51%, liver 50%; p=0.04).

The long-term analysis of FACT-G scores over time are shown in Figure 3, with no differences in total QOL scores, or subscale scores, between arms over time. There were no new grade 2-5 adverse events, and therefore the overall rates of grade ≥ 2 adverse events related to treatment remained at 9% (3/33 patients) in the control arm and 29% (19/66 patients) in the SABR arm (p=0.03), an absolute increase of 20% (95% CI: 5-34%). Notably, as reported previously, there were 3 deaths (4.5%) in the SABR arm that were possibly, probably, or definitely related to treatment.

**Figure 3.**
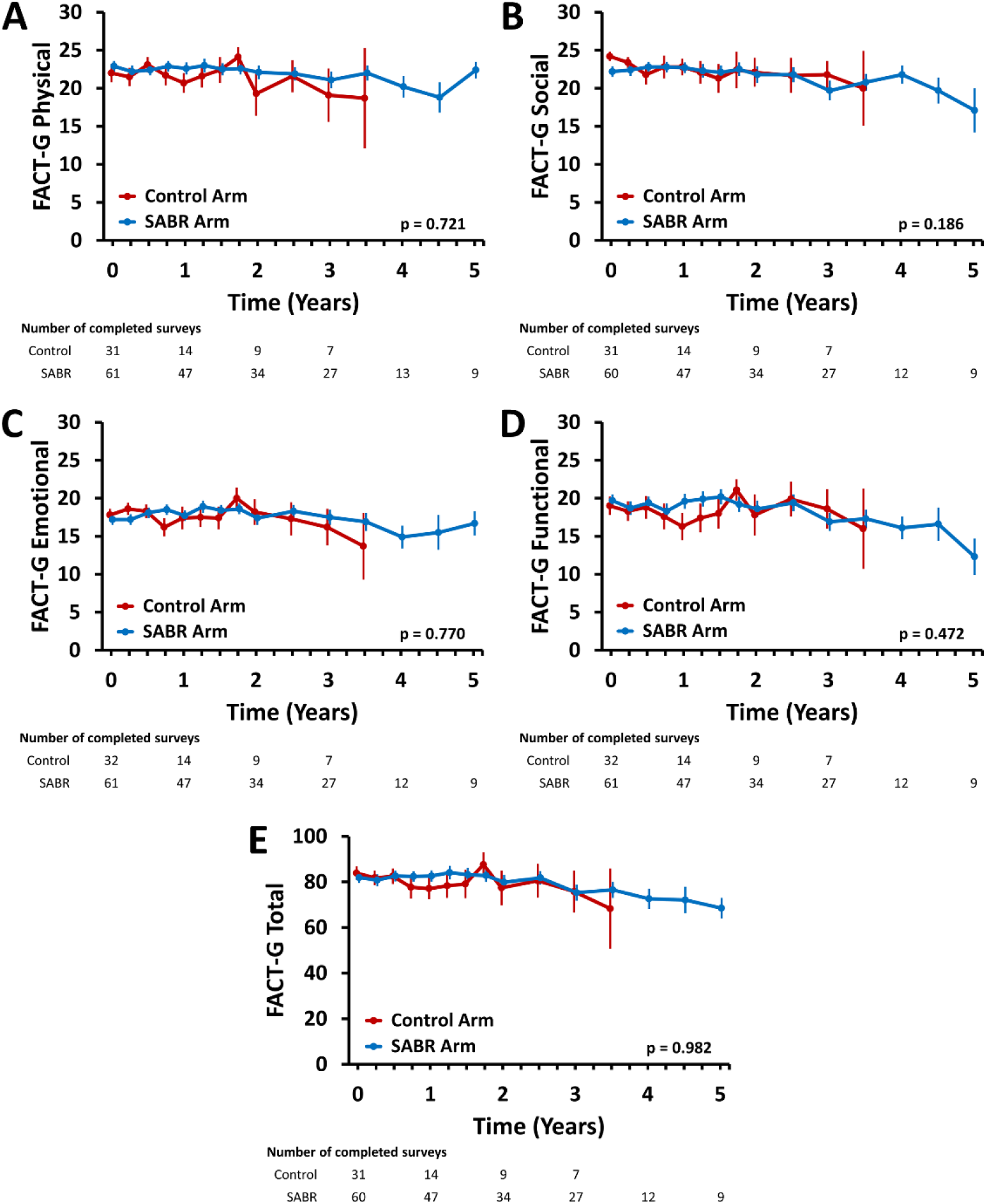
Mean (± standard error) quality of life scores over time, including FACT-G (A) physical well-being, (B) social well-being, (C) emotional well-being, (D) functional well-being, and (E) total score.

The cumulative incidence of new metastases adjusted for death as competing event is shown in Figure 4, with no differences between arms detected (stratified Gray’s test: p=0.57).

**Figure 4.**
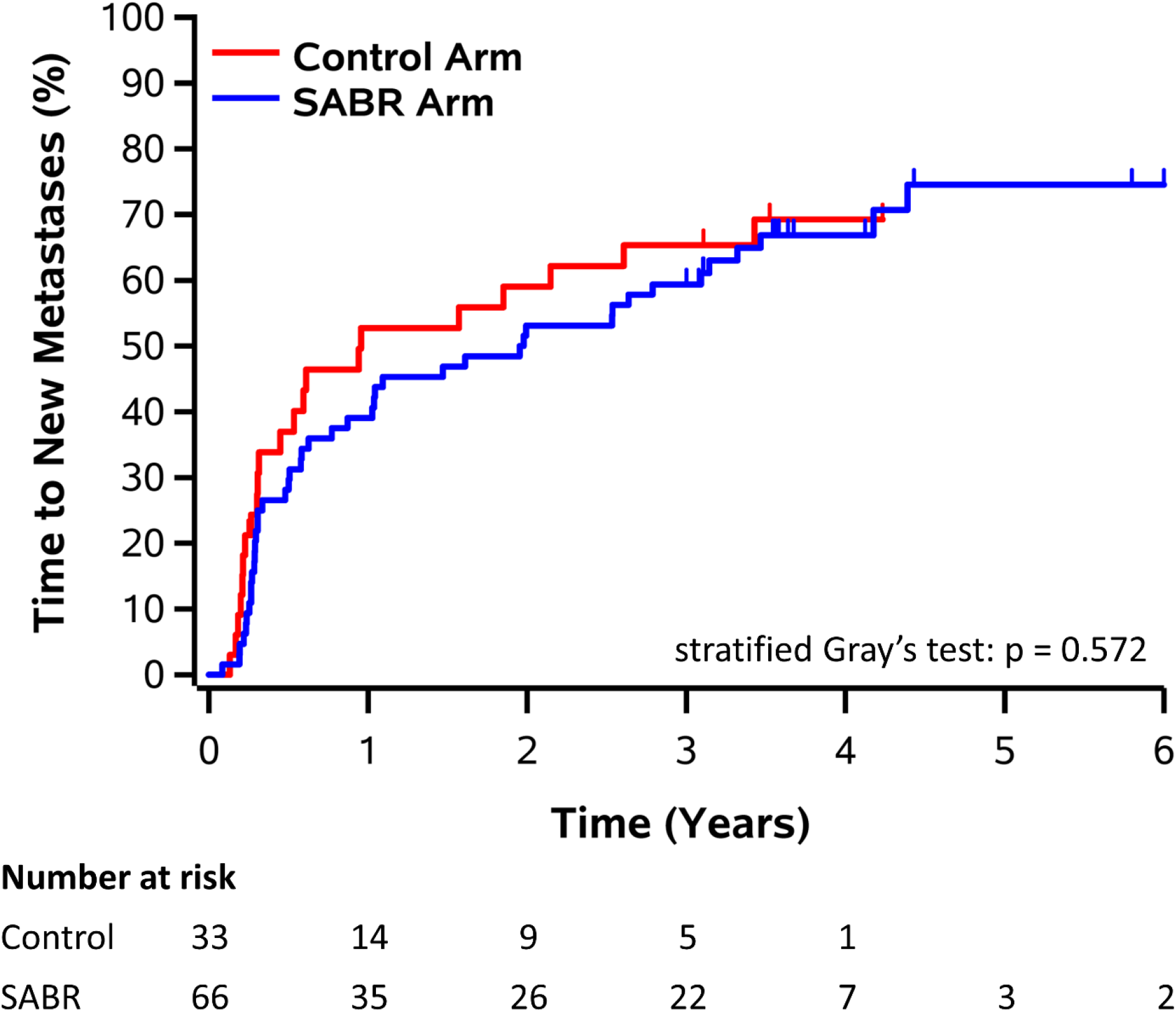
Kaplan-Meier plot for development of new metastases over time based on cumulative incidence function with death as competing event.

## Discussion

At the time of initial publication, SABR-COMET was the first randomized trial demonstrating an impact of any ablative therapy on a primary endpoint of OS in patients with oligometastases. In this long-term analysis, the effects of SABR on OS were larger in magnitude than previously reported, with a median OS benefit of 22 months (compared to 13 months in the original analysis), corresponding to an absolute benefit of 24.6% at 5-years. SABR did not result in a detriment in QOL, and no new safety signals were apparent. The increasing magnitude of benefit over time suggests that long-term follow-up is required for any randomized trials in oligometastatic patients, to fully ascertain the impact of ablative therapies on OS.

It is also apparent from this analysis that most patients with oligometastases have undetectable micrometastases at the time of enrollment, but with close surveillance and further SABR to subsequent developing sites of metastasis, some patients can be successfully treated and again be rendered disease-free. Three lines of evidence support this conclusion: first, there was no significant difference between arms in time to development of new metastases, suggesting that these new metastatic lesions were seeded before SABR was delivered and grew in the months after randomization. Second, a substantial number of long-term survivors (30% of those alive beyond 5 years) required salvage SABR for new metastases. Third, a finding of a comparatively short median PFS benefit (6 months in this trial) in the setting of a longer median OS benefit generally indicates that post-progression treatment is influencing the OS benefit. Since there were no differences in use of systemic therapy between arms, it is likely that post-progression SABR is the main contributing factor to this difference. Taken together, these findings suggest that patients treated with SABR for oligometastases should undergo imaging surveillance with salvage SABR used if safe, as was done in this trial. Further studies are required to determine the optimal imaging surveillance strategy and the maximum number of new lesions treatable with SABR.

Our long-term findings add to a growing body of evidence supporting the use of ablative therapies for oligometastatic cancers. Other phase II trials have suggested benefits of ablative therapies in the setting colorectal cancer liver metastases,^21,22^ in non-small cell lung cancer (NSCLC),^13,15,17^ and in prostate cancer.^16,18^ As a notable exception, the PulMiCC phase III trial failed to show a benefit for surgical resection of pulmonary metastases from colorectal cancers, although the trial closed early and reported on only 21% of target accrual (65 patients).^23^ Overall, however, the preponderance of randomized evidence suggests that patients with oligometastases benefit from ablative therapies, but larger phase III trials, with sufficient power to examine histologic subgroups separately, would be ideal to conclusively prove the survival benefit.

Such phase III trials are underway. SABR-COMET-3 (NCT03862911) and SABR-COMET-10^24^ (NCT03721341) are assessing the impact of SABR on OS in patients with 1-3 and 4-10 metastases respectively, accruing patients with a controlled primary tumor of any solid tumor histology. The CORE trial (NCT02759783) is a phase II/III trial including patients with breast, NSCLC, or prostate histology, with a controlled primary tumor and 1-3 metastatic lesions. Large co-operative group trials specific to lung cancer (NRG-LU002) and breast cancer (NRG-BR002) oligometastases are also underway and accruing well.

Predictive biomarkers would be a major asset to help guide treatment decisions for patients with oligometastases, but currently, no validated biomarkers are available for clinical use. Biomarkers could allow physicians to tailor treatment and surveillance intensity to the risk of further metastatic recurrence. For example, oligometastatic patients predicted to be at high risk of rapid widespread metastatic progression after SABR may be best served by effective systemic therapy rather than SABR (or both treatments in sequence). Efforts to develop biomarkers that are prognostic and predictive are underway as part of ongoing clinical trials; for example, SABR-COMET-3 and SABR-COMET-10 are both collecting samples to assess for circulating biomarkers, including circulating tumor DNA and circulating tumor cells.^24^

The possible toxicities of SABR must be borne in mind for patients and physicians considering treatment. SABR was well-tolerated in the majority of patients, with a rate of grade ≥ 2 toxicity of only 29%. However, the grade 5 toxicity rate of 4.5% (despite strict dose constraints and peer-review of all radiation plans) is higher than reported in other studies. This suggests that SABR delivery should continue to focus on minimization of toxicity, and further studies are needed to determine the optimal SABR doses, balancing the competing considerations of maximizing lesional control while minimizing toxicity.

This trial has limitations that must be considered when interpreting its findings. Many of the limitations were discussed in detail in the original trial report,^14^ including the inclusion of multiple histologies (a common approach in stereotactic radiation trials for metastases). The large majority of patients with prostate cancer were assigned to the SABR arm, but our sensitivity analysis does not suggest that the results are merely due to the allocation of prostate patients: despite the reduced power after excluding prostate patients, a benefit was still demonstrated that would meet the cutoff for a randomized phase II screening trial. Evaluation of local control after SABR is difficult, since focal fibrosis can present as an enlarging mass; this may explain the relatively low local control rates reported for lung lesions using RECIST 1.1.^25^ Patients with 4-5 metastases are under-represented in this trial, which led to the development of separate trials for patients with 1-3 and 4-10 metastases, as described above. This trial was launched before the immunotherapy era, and immunotherapeutic options may change the impact of SABR on long-term outcomes.

In conclusion, with longer-term follow-up, SABR achieved a 22-month median OS benefit in patients with a controlled primary tumor and 1-5 oligometastases. Even with SABR, many patients progress with new metastases, likely due to the presence of occult micrometastatic disease at presentation, but some can be salvaged with repeat SABR. Phase III trials currently underway aim to confirm the OS benefits and to develop biomarkers predictive of benefit with SABR.

## Data Availability

The trial protocol did not include a data sharing plan, and therefore data from the trial will not be shared publicly as sharing was not included in the ethics approvals.

## Acknowledgements

Funded by the Ontario Institute for Cancer Research and a London Regional Cancer Program Catalyst Grant.

## Author Contributions

Trial conception and design: DAP, SS, GBR, ML, CH, BY, BA; Data analysis: AW, DAP Data collection and interpretation: all authors; Initial draft of the manuscript: DAP, SS, AW; Revision of manuscript for critically important content: all authors.

